# Surveying Concerns about the Future in People with Parkinson’s Disease

**DOI:** 10.64898/2026.08.04.26359699

**Authors:** Pooja Gandhi, Li Lin, Theresa Coles, Darby Steiger, Robyn Rapoport, Lana M. Chahine, Connie Marras, Sneha Mantri

**Author notes:** Corresponding author: Sneha Mantri Department of Neurology, Duke University School of Medicine 932 Morreene Rd, Box 3333, Durham, NC 27705, USA.

## Abstract

**Background:** People with Parkinson’s disease (PD) experience substantial psychosocial burden, but the extent of their concern about the future is not well characterized.

**Objective:** The objective of the present study is to characterize concerns about the future among people with PD (PwP), with an emphasis on the clinical and demographic correlates of future-oriented concerns.

**Methods:** A survey in the online Fox Insight study platform asked PwP to rate their degree of concern about the future across seven domains: quality of life, disease progression, healthcare needs, social relationships, financial responsibilities, stigma, and specific symptoms. Relationships among uncertainty and demographic and clinical features, such as age of onset, gender, and disease severity, were examined. Latent class analysis was conducted to identify patterns of fear/uncertainty.

**Results:** Among 3372 respondents, concerns about the future were common and spanned cognitive, functional, social, and symptom-related domains. Concerns about future cognitive impairment, independence, mobility, and disease progression were most prominent. Women and individuals with young-onset PD reported higher levels of concern than other groups. Latent class analysis revealed two clear patterns, including a high-concern subgroup with elevated worry across nearly all domains. Fewer than half of respondents had discussed these concerns with a healthcare professional.

**Conclusion:** Future-related concerns are common among people with PD but is not routinely explored in clinical care. Greater attention to these concerns, especially for young-onset individuals and women, may help clinicians offer more timely and supportive guidance.

**Plain Language Summary:** Many people with PD worry about their future, including uncertainty around cognition, independence, and disease progression. These concerns appear to be greatest in women and people with young-onset PD, yet few report discussing their concerns with their clinicians. Anticipatory guidance from clinicians about future-related worries may mitigate uncertainty-related psychosocial distress.

---

Parkinson’s disease (PD) is a progressively debilitating neurological disorder projected to affect over 12 million individuals globally by 2040.[1] It significantly disrupts patients’ daily lives, presenting a range of challenges that vary across the disease trajectory, from diagnosis through the later stages.[2] While some difficulties are confined to particular phases of the illness, others persist throughout its course.[3] PD impacts numerous aspects of psychosocial functioning, including cognition,[4] mood,[5] social relationships,[6,7] psychological well-being,[8,9] and communication.[10,11]

The substantial variability in individual disease presentations complicates clinicians’ ability to prognosticate the course of illness for any given patient[3,12]. This unpredictability contributes to heightened psychosocial burden, disease-related anxiety, and emotional distress.[13] Similar patterns of uncertainty and psychological impact have been observed in other chronic illnesses, such as asthma,[14] inflammatory arthritis,[15] and cancer.[16]

Prior qualitative research has identified several domains of fear commonly experienced by people with Parkinson’s (PwP). Specifically, an analysis of patient-reported “most bothersome problems” highlighted two major sources of fear: fear of cognitive decline and fear of falling.[17] These fears not only contribute to uncertainty but also significantly reduce quality of life for both PwP and their care partners.[18–20] Importantly, these concerns often resist standard treatment approaches and are frequently regarded as markers of disease progression.[21,22]

More recently, Trahair et al. (2025) expanded the understanding of fear and uncertainty in PD through a qualitative analysis of narratives shared by PwP.[23] Their findings highlighted multiple future-oriented fears but did not quantify their prevalence or relative importance. Building on this work, the present study seeks to further explore concerns about the future among PwP. We aimed to survey the breadth, relative frequency and intensity of concerns about the future reported by PwP.

Specifically, the study objectives were (1) to describe categories of concern by age of onset, gender identity, and race/ethnicity; and (2) to examine how demographic and clinical factors, including mood, education level, living situation, and disease severity, are associated with the intensity of concern experienced.

## Methods

This study was phase 3 of a 3-part mixed-methods investigation. Phases 1 and 2 consisted of exploratory and semi-structured interviews with people living with Parkinson’s disease (PwP) as described in detail in Trahair et al. (2025). Results of phases 1 and 2 were used to develop the survey administered in phase 3. The study was approved by the WCG-IRB (Protocol 20235671) and informed consent was obtained from all participants.

Briefly, interviews conducted between April and July 2024 identified a broad set of perceived and anticipated concerns about the future, refined terminology, and informed the preliminary item pool for the survey. Insights from these phases guided the development and cognitive testing of the final questionnaire. Each survey question asked “How concerned are you about each of the following when you think about your future with PD?”, as pertains to specific domains: quality of life (5 items), disease progression (6 items), social impact (3 items), financial concerns (4 items), and specific symptoms (6 items). For each item, participants rated how concerned they were about the issue when thinking about their future with PD using a 5-point Likert scale with response options ranging from “not at all concerned” to “extremely concerned.” After completing the Likert ratings, participants were asked to rank-order the issues about which they felt most uncertain or fearful (Likert ratings of 4 or 5) and whether they had shared these top concerns with others.

Phase 3, reported here, involved administering the final questionnaire to individuals enrolled in Fox Insight, a longitudinal, online research platform launched by MJFF in 2017. Fox Insight collects self-reported data from PwP at regular intervals using web-based questionnaires and is designed to support large-scale, remote participation.[24] Participants complete surveys through a secure online portal. Fox Insight data has previously been used to analyze most bothersome symptoms,[25,26] including analysis of fear as a discrete contributor to psychosocial distress.[27] For this study, the survey about concerns for the future was distributed as an optional questionnaire within the Fox Insight platform, and responses were captured directly through the study’s digital infrastructure.

### Statistical analysis

Descriptive statistics were computed for all study variables. For continuous variables, we reported the mean, median, standard deviation, and interquartile range. Categorical variables were summarized using frequencies and proportions.

Depressive symptoms were assessed using the 15-item Geriatric Depression Scale (GDS-15),[28] a validated self-report screening instrument with scores ranging from 0 to 15, where higher scores indicate greater depressive symptom burden. A score of ≥5 was used to indicate clinically significant depressive symptoms. Clinically anchored disease severity was categorized using thresholds from the neuronal alpha-synuclein integrated staging system.[29]

To examine the distribution of each Likert-scale item, we calculated both the frequency of response options and the mean and standard deviation of scores. Group differences were assessed using Wilcoxon rank-sum tests for statistical significance and effect sizes for magnitudes.[30]

Areas of concern were ranked by average item response scores to identify the most prominent issues reported by patients with Parkinson’s disease.

While item-level group comparisons were examined as secondary analyses, our primary analytic approach was latent class analysis, which identifies subgroups of participants based on patterns across all survey items simultaneously. This person-centered approach provides a more clinically interpretable summary than multiple single-item comparisons, which can yield statistically significant but clinically small differences.

Latent class Analysis (LCA) was conducted to identify distinct subgroups (related areas of concern) within the sample based on the Likert-scale indicators. Model fit was assessed using multiple criteria, including the Bayesian Information Criterion (BIC), Akaike Information Criterion (AIC), entropy, and likelihood ratio tests. The optimal number of latent classes was selected based on statistical fit and interpretability. For each individual, the model calculated the probability of belonging to each subgroup given their observed data and assigned each person to the subgroup with the highest posterior probability. Mean profiles of the identified subgroups were visualized to aid interpretation.

To compare demographic and clinical characteristics across latent subgroups, we used Wilcoxon rank-sum tests for continuous variables and chi-square tests for categorical variables. Contingency tables were employed to explore whether patients with higher levels of concern were more likely to discuss their concerns with others. Additionally, we examined the terminology patients used to describe uncertainty.

Data used in the preparation of this article were obtained from the Fox Insight database (https://foxinsight-info.michaeljfox.org/insight/explore/insight.jsp) on 15 JUL 2025. Fox Insight is an online longitudinal cohort that primarily enrolls participants from the United States, although individuals from other countries are also eligible to participate. Participation in Fox Insight is voluntary and participants are not remunerated for completing surveys. Because participation is remote and web-based, the cohort reflects a geographically distributed sample rather than recruitment from specific clinical centers. For up to date information on the study, visit https://foxinsight-info.michaeljfox.org/insight/explore/insight.jsp. LCA was performed using Mplus Version 8.11. Data visualization was conducted in R Version 4.1.2, and all other statistical analyses were carried out using SAS Version 9.4

## Results

### Demographic and clinical characteristics

Between February 4 and July 15, 2025, 3387 individuals accessed the survey in Fox Insight. Fifteen respondents who did not report a diagnosis of Parkinson’s disease (PD) were excluded, resulting in 3372 completed surveys (99.6% completion rate). (Table 1). The mean age of respondents was 69.3 years (SD 8.9). The mean age at diagnosis was 60.4 years, and 14.4% had young-onset PD, defined as diagnosis at or before age 50 years. Just over half of participants identified as women (51.2 percent), 47.7 percent as men, and the remainder as another gender or preferred not to answer. The cohort was predominantly non-Hispanic White (91.7 percent). Educational attainment was high, with more than 70 percent reporting a bachelor’s degree or higher, and most respondents reported living with a spouse or family member. Clinically anchored disease severity, based on UPDRS-II staging available through Fox Insight (26), was distributed primarily across stages 3 (very mild symptoms with no functional impact) and 4 (mild symptoms with mild functional impact). Respondents were more likely than the broader Fox Insight cohort to report a history of depression or anxiety.

**Table 1.** Demographic and Clinical Characteristics of Survey Respondents (N=3372)

| Characteristic | Mean (SD) or N (%) |
| --- | --- |
| Age at PD onset (Mean, SD) | 60.4 (9.3) |
| Age at time of survey (Mean, SD) | 69.3 (8.9) |
| Young-Onset Parkinson disease (YOPD; age at diagnosis <50) (N %) | 475 (14.4%) |
| Race / Ethnicity (n, %), Check all that apply |  |
| Non-Hispanic White or Caucasian | 3034 (91.7%) |
| Black or African American | 13 (0.4%) |
| American Indian or Alaska Native | 23 (0.7%) |
| Asian | 45 (1.4%) |
| Native Hawaiian or Other Pacific Islander | 4 (0.1%) |
| Hispanic/Latino | 126 (3.8%) |
| Prefer not to answer | 90 (2.7%) |
| Missing or unknown | 63 (1.9%) |
| Gender (n, %) |  |
| Male | 1578 (46.80%) |
| Female | 1694 (50.24%) |
| Unknown | 100 (2.97%) |
| Geriatric Depression Scale (GDS, mean, SD) | 3.7 (3.6) |
| GDS ≥ 5 (n, %) | 1032 (31.0%) |
| Education level (n, %) |  |
| Less than a high school degree | 24 (0.73%) |
| High school degree | 197 (5.95%) |
| Some college | 478 (14.45%) |
| Associate's degree | 276 (8.34%) |
| Bachelor's degree | 988 (29.86%) |
| Master's degree | 908 (27.44%) |
| Professional school degree | 233 (7.04%) |
| Doctorate degree | 191 (5.77%) |
| Prefer not to answer | 14 (0.42%) |
| Missing or unknown | 63 (1.9%) |
| Clinically anchored disease severity |  |
| Stage 2, subtle clinical symptoms without functional impairment | 269 (8.05%) |
| Stage 3, clinical symptoms with slight functional impairment | 1725 (51.62%) |
| Stage 4, clinical symptoms with mild functional impairment | 1144 (34.23%) |
| Stage 5, clinical symptoms with moderate functional impairment | 182 (5.45%) |
| Stage 6, clinical symptoms with severe functional impairment | 22 (0.66%) |
| Missing or unknown | 30 (0.9%) |
| Living situation |  |
| Live alone | 335 (14.4%) |
| Live with other family members (spouse/partner, child, other family) | 1962 (84.1%) |
| Live with paid caregiver/in nursing home/assisted living | 25 (1.1%) |
| Other living situation | 11 (0.5%) |
| Missing or unknown | 1039 (30.8%) |

### Descriptive patterns of concern

Participants reported concerns related to their future across all domains queried (Table 2). The highest mean levels of concern were observed for cognitive impairment, loss of independence, loss of mobility, and overall disease progression. Concerns related to personal care needs and the anticipated impact of PD on family members also ranked among the higher mean scores. In contrast, lower mean levels of concern were observed for stigma, invasive treatments, individualized treatment access, and issues involving death and dying.

**Table 2.** Top five highest-ranked concerns.

| DOMAIN | Uncertainty Item | Mean | Lower 95% CL for Mean | Upper 95% CL for Mean |
| --- | --- | --- | --- | --- |
| SYMPTOMS | Cognitive impairment, such as loss of memory, difficulty finding words, confusion, dementia, multi-tasking, or other cognitive impairment issues | 3.72 | 3.68 | 3.76 |
| QUALITY OF LIFE | Loss of independence, such as losing the ability to drive, to make your own decisions, or other ways you might lose independence | 3.57 | 3.53 | 3.61 |
| DISEASE PROGRESSION AND CARE NEEDS | Disease progression, such as worsening of your PD symptoms, uncertainty about how the disease will progress, long term effectiveness of medications, or other concerns around disease progression | 3.54 | 3.51 | 3.58 |
| QUALITY OF LIFE | Loss of mobility, such as losing the ability to exercise, needing a walker or a wheelchair, or other loss of mobility | 3.50 | 3.47 | 3.54 |
| RELATIONSHIPS SOCIAL IMPACT | Impact on your family, such as family or close friends, potentially being a burden on others, impact on your relationships, physical intimacy with your spouse or partner, or other impacts on your family | 3.45 | 3.41 | 3.48 |

### Subgroup differences and highest concerning domains

Latent class analysis identified two distinct groups based on patterns of responses (Figure 1). Class 1 represented 43.6% of the sample (n = 1,472) and was characterized by consistently lower concern across all domains, with mean scores generally ranging from approximately 1.9 to 2.7. In contrast, Class 2 comprised 56.4% of respondents (n = 1,900) and showed markedly higher levels of concern, with means spanning roughly 3.3 to 4.4 across indicators. The largest class differences were observed for cognitive symptoms (2.89 vs. 4.36), independence (2.69 vs. 4.25), mobility (2.64 vs. 4.17), disease progression (2.74 vs. 4.16), autonomic symptoms (2.58 vs. 3.97), and impact on family members (2.63 vs. 4.07), all p < 0.001. These separations indicate that more than half of respondents fell into a profile characterized by elevated concern across multiple domains.

**Figure 1.**
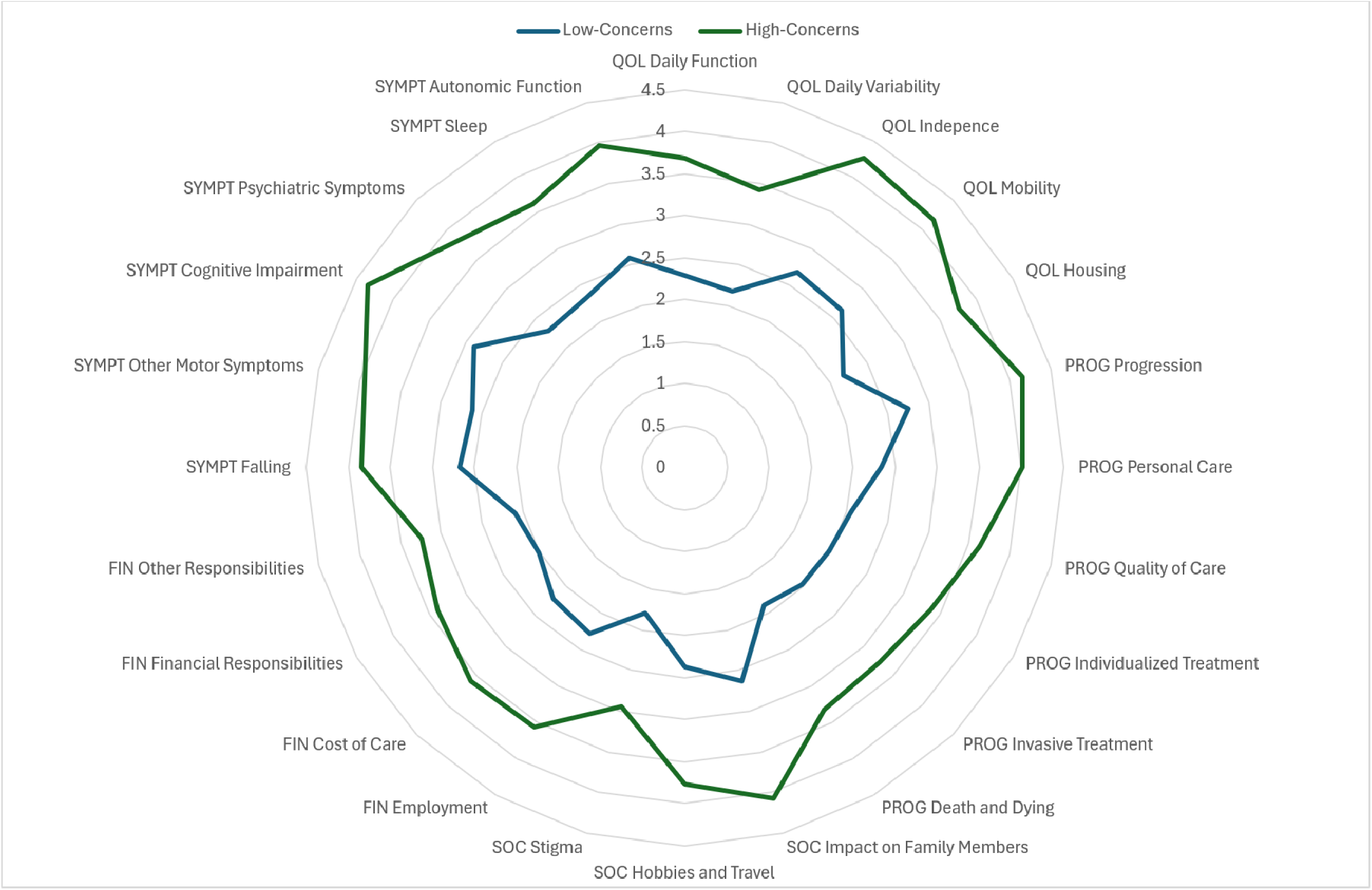
Radar Plot of mean concern ratings by item of High and Low Concern Groups Mean concern scores (Likert scale 1–5) are shown for each item across the two latent classes identified by latent class analysis. Domain abbreviations: QOL = quality of life; PROG = disease progression and care needs; SOC = social impact; FIN = financial concerns; SYMPT = symptoms.

Class membership was associated with several demographic and clinical characteristics (Table 3). Participants in the high-concern class were more likely than those in the low-concern class to be women and to have young-onset PD. The high-concern class also had a higher prevalence of clinically significant depressive symptoms (GDS ≥5) and greater clinically anchored disease severity as measured by responses to UPDRS Part II. Differences were also observed by educational attainment and living situation.

**Table 3.** Severity of concerns within clinical and demographic groups.

| Variable | Level or Statistic | Low Concerns (Class 1)<br>(N=1472) | High Concerns (Class 2)<br>(N=1900) | P-value+ |
| --- | --- | --- | --- | --- |
| Young-Onset Parkinson's Disease (AgeatDx <= 50) | Missing | 23 (1.56%) | 45 (2.37%) | **0.0009** |
|  | No | 1274 (87.92%) | 1555 (83.83%) |  |
|  | Yes | 175 (12.08%) | 300 (16.17%) |  |
| Race and ethnicity | Missing | 27 (1.83%) | 36 (1.89%) | 0.8007 |
|  | Non-Hispanic White only | 1330 (92.04%) | 1704 (91.42%) |  |
|  | All others | 78 (5.40%) | 107 (5.74%) |  |
|  | Prefer not to answer on race and/or ethnicity | 37 (2.56%) | 53 (2.84%) |  |
| Gender | Male | 786 (53.40%) | 792 (41.68%) | **<.0001** |
|  | Female | 645 (43.82%) | 1049 (55.21%) |  |
|  | Other, unknown, or missing | 41 (2.79%) | 59 (3.11%) |  |
| Geriatric Depression Scale Score | Missing | 15 (1.02%) | 26 (1.37%) | <.0001 |
|  | GDS < 5 | 1206 (82.77%) | 1093 (58.32%) |  |
|  | GDS ≥5 | 251 (17.23%) | 781 (41.68%) |  |
|  | Mean (SD) | 2.5 (2.6) | 4.6 (4.0) | < 0.001 |
| Education level | Missing | 27 (1.83%) | 36 (1.89%) | 0.0024 |
|  | Some HS | 6 (0.42%) | 18 (0.97%) |  |
|  | HS diploma | 82 (5.67%) | 115 (6.17%) |  |
|  | Some college, no degree | 188 (13.01%) | 290 (15.56%) |  |
|  | Associate's degree | 98 (6.78%) | 178 (9.55%) |  |
|  | Bachelors degree | 432 (29.90%) | 556 (29.83%) |  |
|  | Masters degree | 431 (29.83%) | 477 (25.59%) |  |
|  | Professional degree | 114 (7.89%) | 119 (6.38%) |  |
|  | Doctorate degree | 89 (6.16%) | 102 (5.47%) |  |
|  | Prefer not to answer | 5 (0.35%) | 9 (0.48%) |  |
| Clinically anchored disease severity | Missing | 9 (0.61%) | 21 (1.11%) | <.0001 |
|  | 2 | 171 (11.69%) | 98 (5.22%) |  |
|  | 3 | 851 (58.17%) | 874 (46.51%) |  |
|  | 4 | 408 (27.89%) | 736 (39.17%) |  |
|  | 5 | 30 (2.05%) | 152 (8.09%) |  |
|  | 6 | 3 (0.21%) | 19 (1.01%) |  |
| Live Alone | Missing | 411 (27.92%) | 628 (33.05%) | 0.0185 |
|  | Live alone | 153 (14.42%) | 182 (14.31%) |  |
|  | Live with other family members (spouse/partner, adult child, minor child, other family) | 901 (84.92%) | 1061 (83.41%) |  |
|  | Live with paid caregiver/in nursing home/assisted living | 5 (0.47%) | 20 (1.57%) |  |
|  | Live with other | 2 (0.19%) | 9 (0.71%) |  |

+ Note: P-values were calculated by comparing non-missing row values only; these percents sum to 100%. The percent of missing row values is informative and therefore also presented here for convenience.

In secondary item-level analyses, women and individuals with young-onset PD reported higher mean concern ratings across multiple domains, including cognition, mobility, and several non-motor symptom concerns. Item-level comparisons by gender, age of onset, and race/ethnicity are provided in Supplementary Table S1.

Overall, 35.5% of participants reported discussing their concerns with their main PD healthcare provider (Table 4), and 17.0% reported not having talked to anyone about these issues. Of those who had sought support regarding fears and uncertainties, most had spoken with their spouse (n=2212, 65.6%), other family members (n=1240, 36.8%), or friends (n=1191, 35.3%). This analysis was descriptive; differences in disclosure patterns across demographic groups were not examined.

**Table 4.** Individuals with whom respondents discussed concerns about the future related to PD.

| **Variable** | **Count** | **Percentage** |
| --- | --- | --- |
| Spouse | 2212 | 65.6% |
| Other family members | 1240 | 36.8% |
| Friends | 1191 | 35.3% |
| Support groups/other people with PD | 803 | 23.8% |
| Your main healthcare provider for PD | 1198 | 35.5% |
| Other healthcare providers | 434 | 12.9% |
| A counselor or therapist | 456 | 13.5% |
| Clergy or religious leaders | 88 | 2.6% |
| I typically do not talk with anyone about these issues | 573 | 17.0% |
| Other | 64 | 1.9% |
| Prefer not to answer | 13 | 0.4% |

## Discussion

In this large online sample of PwP, uncertainty about the future was a common and wide-ranging concern. Participants endorsed substantial worry across physical, cognitive, social, and financial domains, with the highest levels of concern centered on cognitive impairment, loss of independence, and loss of mobility. In earlier qualitative work, PwP described fears related to declining function, future care needs, and changes in identity and social roles.[23] The present study extends that work by quantifying the prevalence and relative intensity of these concerns in a large, well-characterized cohort.[24] The age distribution and gender balance of this sample are broadly consistent with epidemiologic descriptions of PD populations;[31] however, participants were more highly educated and less racially diverse than many community-based cohorts, which may limit generalizability.

Latent class analysis identified two distinct profiles of uncertainty: a lower-concern group and a higher-concern group. More than half of respondents fell into the higher-concern class, characterized by elevated worry across most domains. The largest differences between classes involved cognitive decline, independence, mobility, disease progression, autonomic symptoms, and the anticipated impact on family members. These findings reinforce the central role of these concerns in shaping how PwP think about their future.

A notable finding was the higher burden of uncertainty among women and individuals with young-onset PD. Women were more likely to belong to the higher-concern class and reported greater concern across multiple domains, including cognition, mobility, sleep, psychiatric symptoms, and family impact. Individuals with young-onset PD were similarly more likely to fall into the high-concern class and expressed elevated worry about employment, finances, long-term independence, and family responsibilities, reflecting the distinct life-course challenges associated with earlier diagnosis. Together, these findings suggest that young-onset women may represent a subgroup with particularly elevated uncertainty.

Clinical characteristics were also associated with higher uncertainty. Participants with depressive symptoms were more likely to fall into the high-concern class, consistent with prior evidence linking mood and illness-related worry in PD (5). Greater disease severity, lower educational attainment, and living alone were similarly associated with higher levels of concern. These findings highlight groups who may benefit from more structured anticipatory guidance.

Although uncertainty was common, fewer than half of respondents reported discussing these concerns with a clinician. This aligns with previous qualitative findings showing that patients often hesitate to raise future-oriented fears, and that clinical encounters tend to focus primarily on motor symptom management.[23] The disconnect between patient experience and clinical discussion suggests an opportunity for more proactive inquiry about concerns related to cognition, functional independence, family impact, and long-term planning. These conversations may be particularly important for patients at higher risk of elevated uncertainty.

This study has several strengths, including the large sample size, broad range of uncertainty domains assessed, and the use of latent class analysis to identify distinct patterns of concern. However, several limitations should be considered. This survey was developed from semi-structured interviews with PwP but is not a validated scale. As participation in this survey was optional, the sample may overrepresent individuals with higher psychosocial burden, including those with depression or anxiety. The cross-sectional design limits our ability to characterize how uncertainty evolves over time. The racial and ethnic composition of the sample was largely homogeneous, with a predominance of white and highly educated individuals with a narrow range of disease severity which may limit generalizability to more diverse populations, particularly as several of the uncertainties assessed are culturally mediated (e.g. hobbies, stigma, and relationships with family). Finally, although the measure captured a wide range of concerns, certain domains such as caregiver burden, existential concerns, or specific challenges faced by underrepresented groups were not directly assessed.

Concern about the future is a common and multifaceted experience for people living with Parkinson’s disease. A substantial proportion of PwP fall into a profile marked by elevated concern across domains, yet many do not discuss these concerns in clinical care. Identifying patients at higher risk of elevated uncertainty may allow for more proactive and supportive conversations about future expectations and planning. Providing accessible educational or counseling resources that address common areas of uncertainty may also help patients and families develop strategies for coping with these concerns. Continued longitudinal work is needed to understand how these concerns evolve and how best to address them in routine care.

## Data Availability

All data produced are available online at FoxDen.michaeljfox.org

https://foxden.michaeljfox.org/insight/explore/insight.jsp

## Acknowledgements

The Fox Insight Study (FI) is funded by The Michael J. Fox Foundation for Parkinson’s Research. We would like to thank the Parkinson’s community for participating in this study to make this research possible.

## Author Roles

PG: 1A, 1B, 2C, 3A

LL: 2A, 2B, 3B

TC: 2A, 2C, 3B

DS: 3B

RR: 3B

LMC: 1A, 1B, 3B

CM: 1A, 1B, 3B

SM: 1A, 1B, 2C, 3B

## Statements and Declarations

### Ethical Considerations

This study was approved by the WCG IRB (Protocol 20235671).

### Consent to Participate

Informed consent was obtained electronically from all participants via the FoxDEN study platform.

### Consent for Publication

Not applicable

### Data availability statement

The data used in this study is available upon registration with the FoxDEN study platform (https://foxden.michaeljfox.org/insight/explore/insight.jsp)

---

**Supplemental Table 1.**
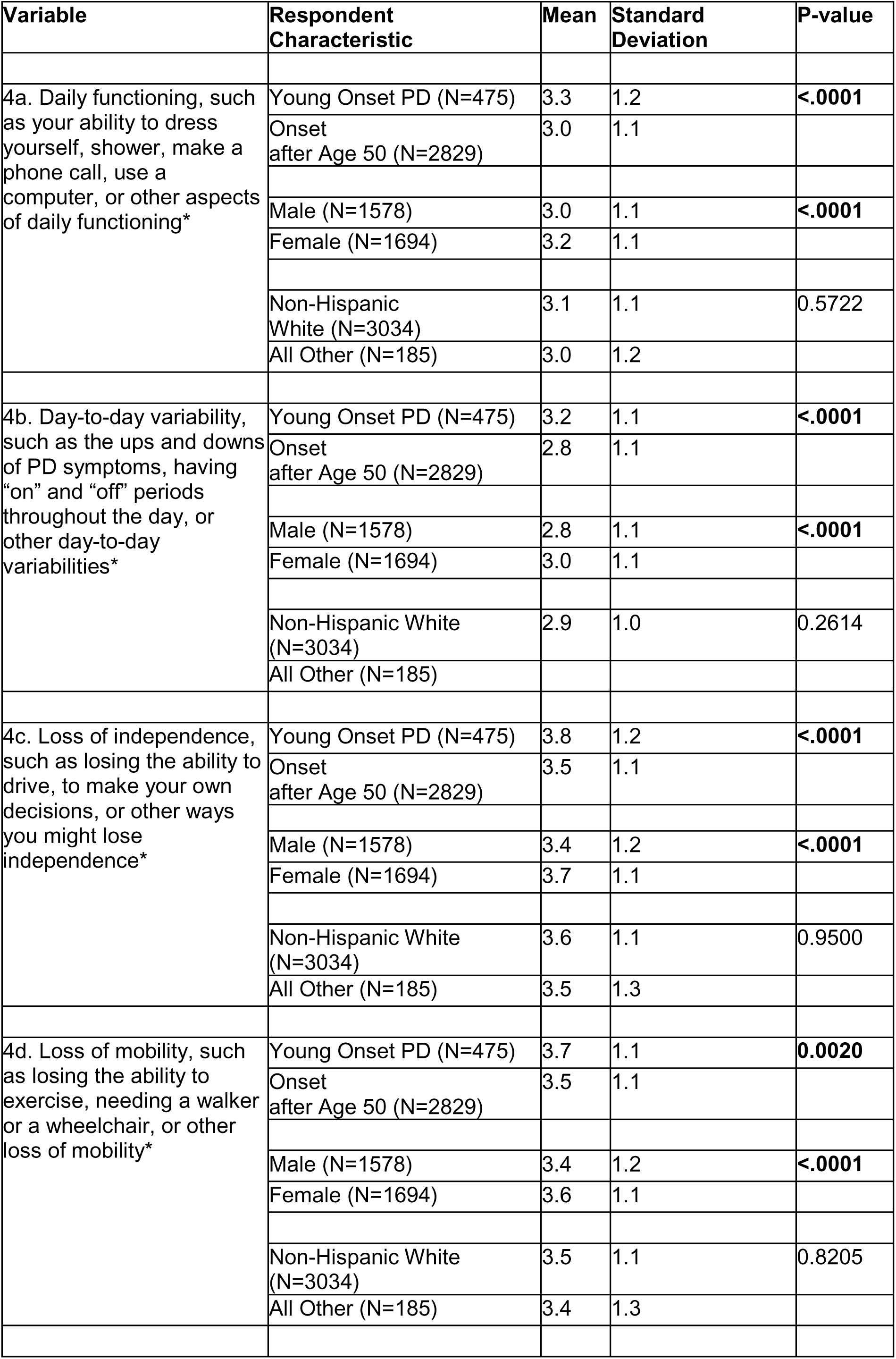
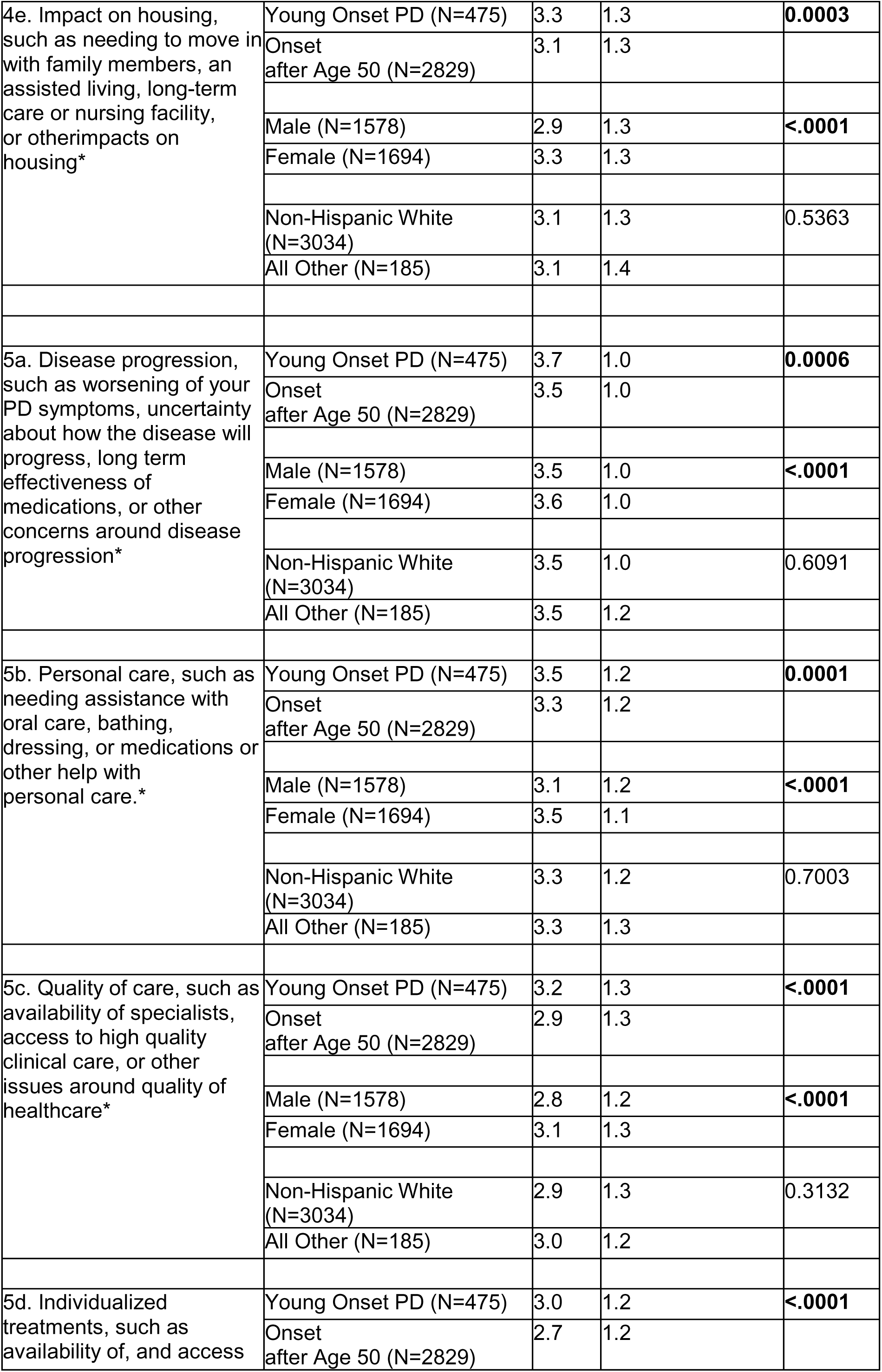
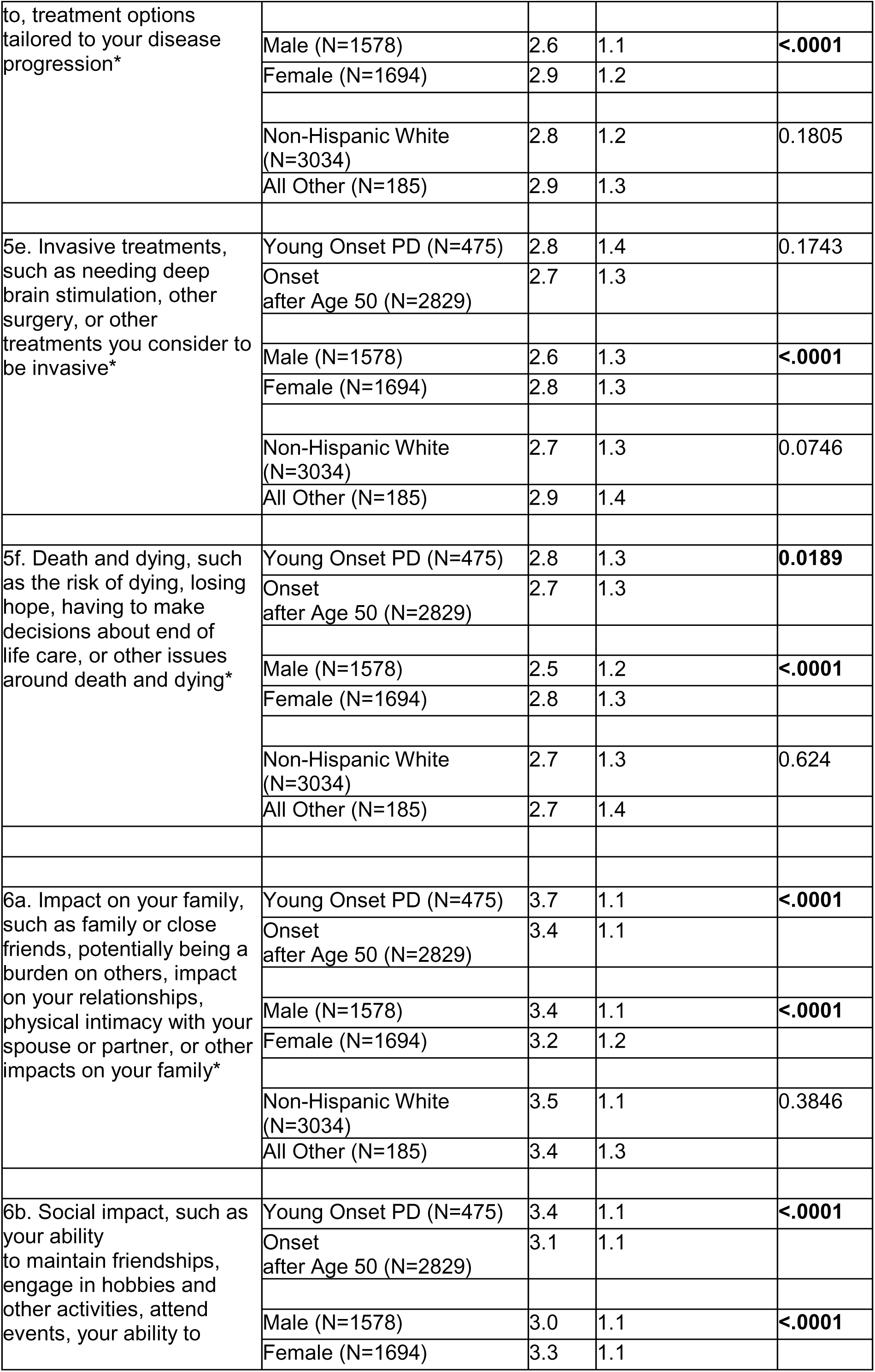
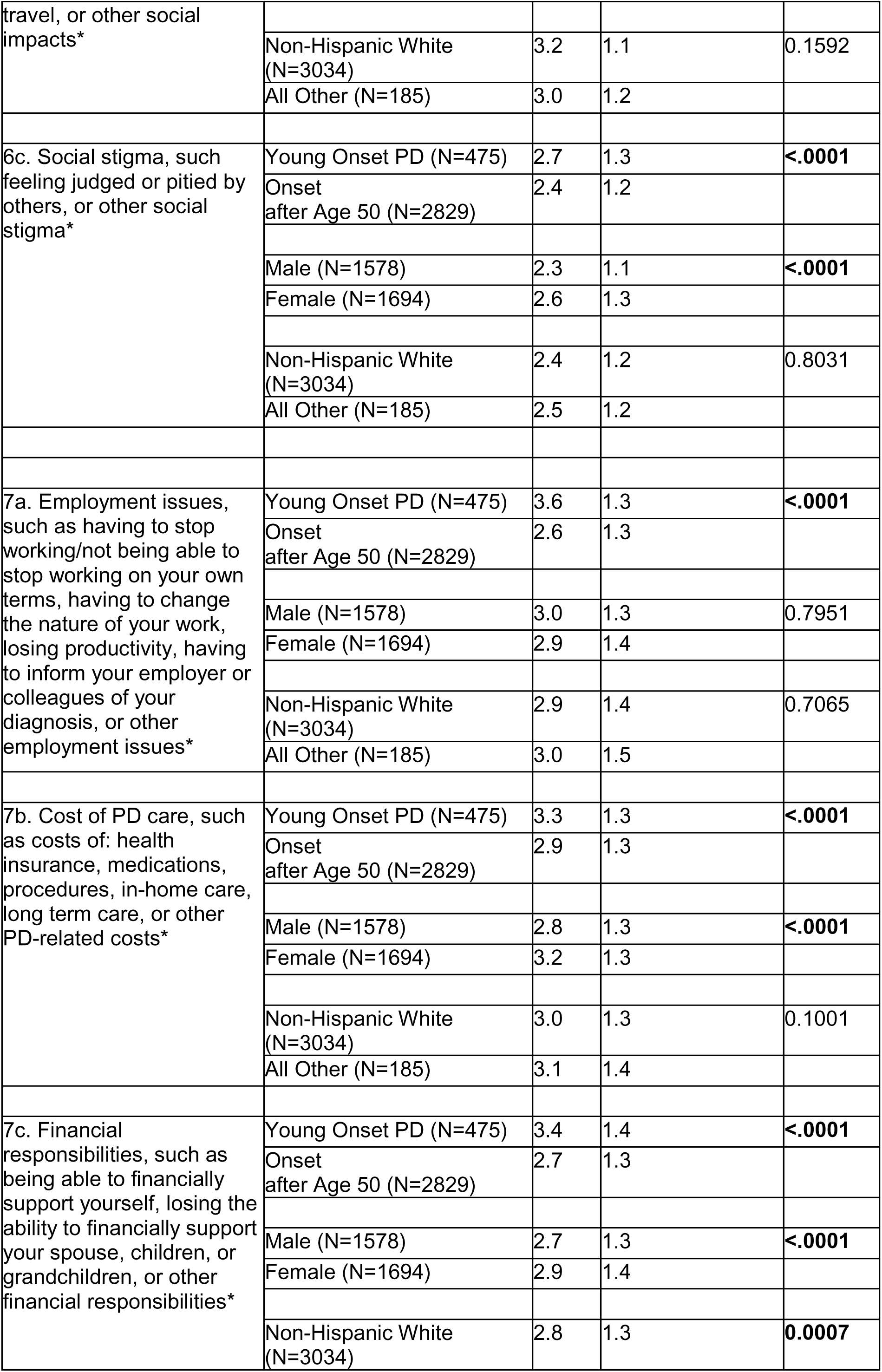
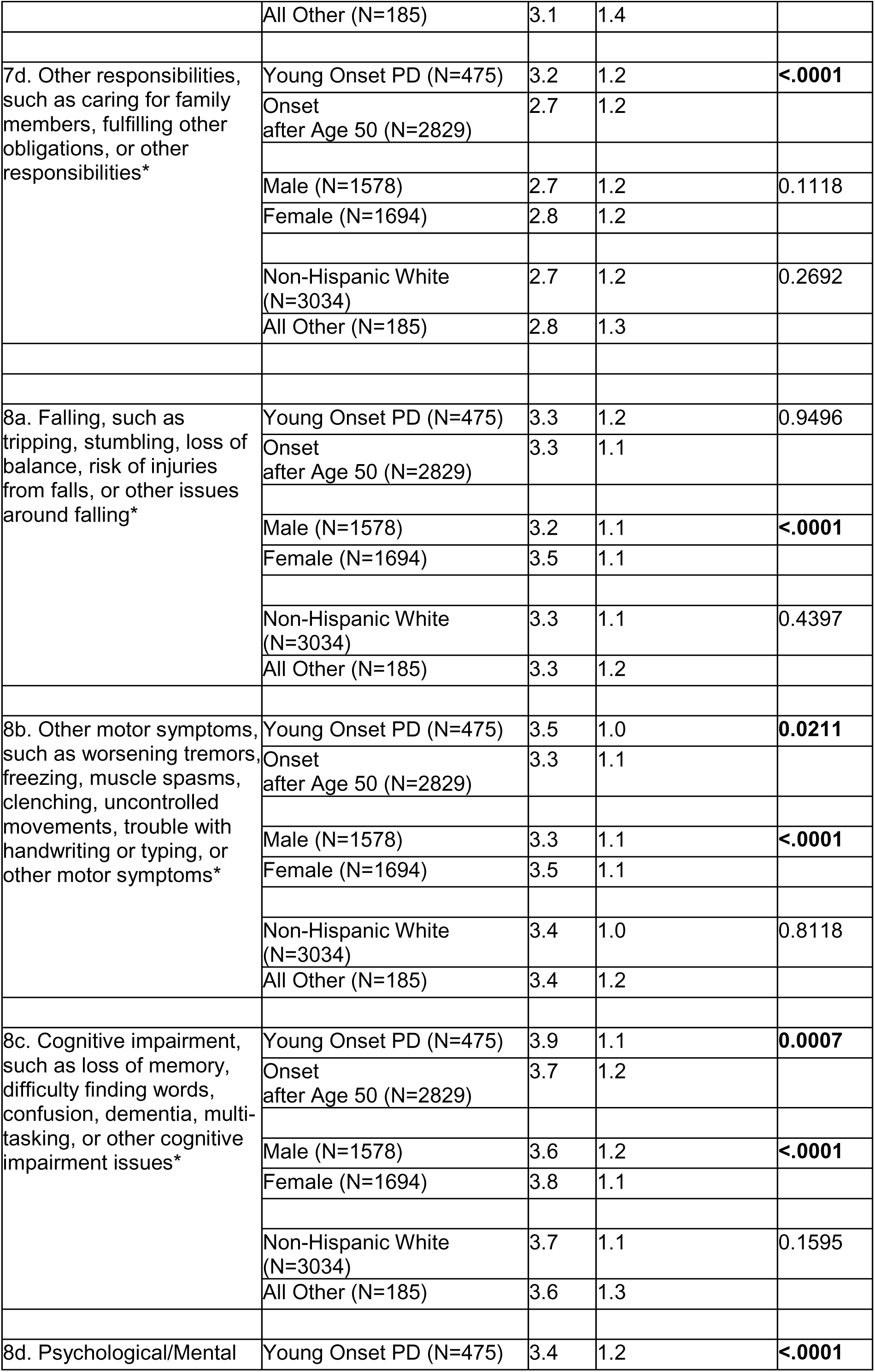
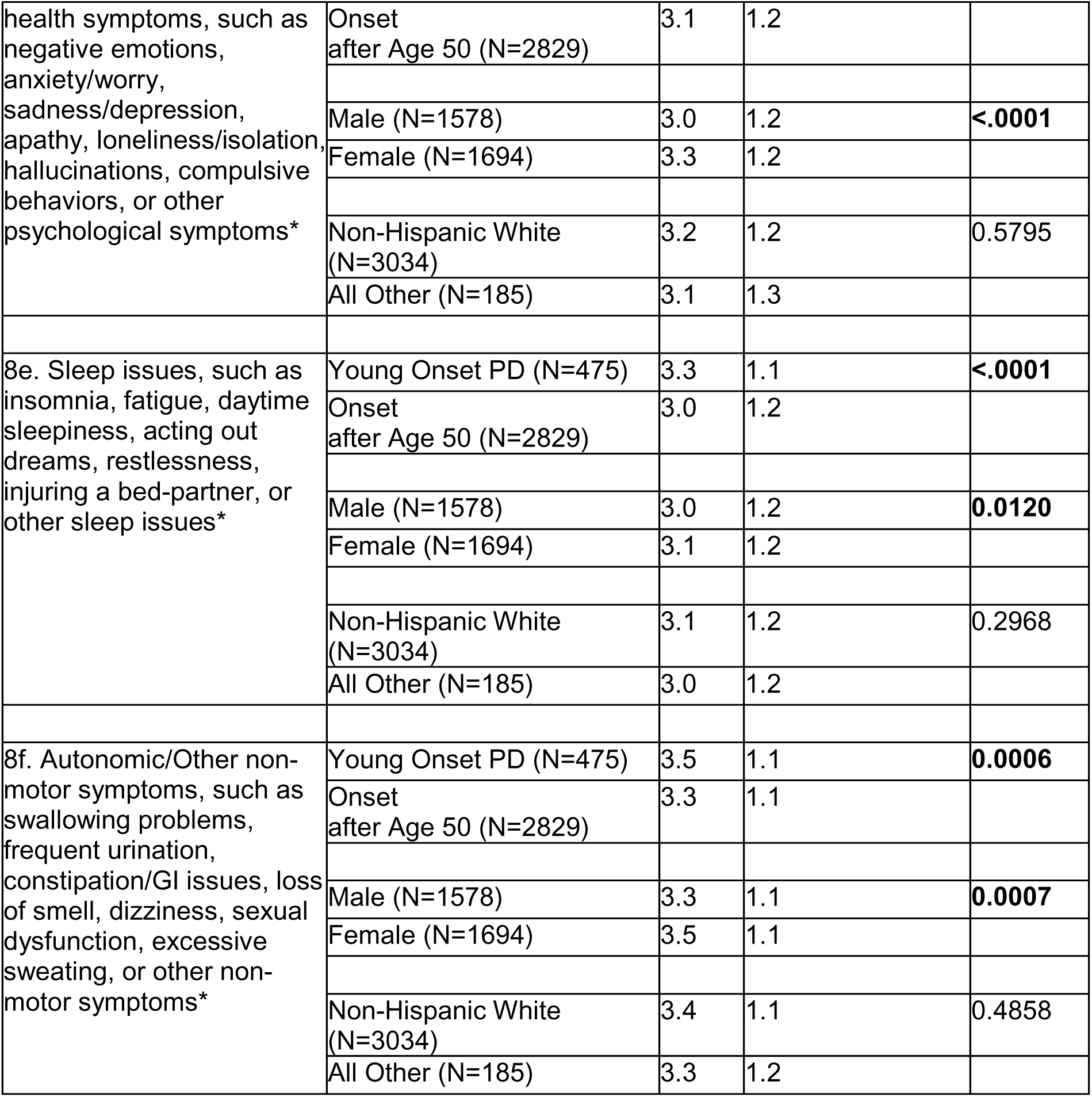
Item-level uncertainty ratings, by age of onset, gender, and race/ethnicity.

